# Screen-Detected and Diagnostic Breast Cancers Show Distinct Treatment Pathways and Quality Indicator Performance

**DOI:** 10.64898/2026.07.13.26357901

**Authors:** Zuzana Bielčiková, Aleš Tichopád, Marian Rybář, Katarína Petráková, Martin Rožánek, Karla Mothejlová, Ladislav Dušek, Gleb Donin

## Abstract

Population-based mammography screening improves breast cancer outcomes, but its impact on real-world treatment pathways and quality indicators (QIs) remains incompletely described. We conducted a retrospective nationwide cohort study using linked data from the Czech National Cancer Registry and the National Registry of Reimbursed Health Services. Women aged ≥18 years with a first breast cancer diagnosis between 2017 and 2024 were classified as screen-detected (SCR) or diagnostically-detected (DIG) according to the imaging modality preceding histological verification. Outcomes included stage distribution, untreated cases, first-line treatment, main treatment modality, time to treatment, multidisciplinary team discussion (MDT), centralization to Comprehensive Cancer Centres (COCs), and survival patterns. The verified cohort included 47,648 women: 26,817 SCR cases (56.3 %) and 20,831 DIG cases (43.7 %). In this nationwide analysis, SCR breast cancer was associated with earlier stage at diagnosis and better survival patterns, but also with longer time to treatment and longer time to MDT discussion than DIG-detected disease. Although treatment rates were high and centralization improved over time, substantial regional variation persisted in care pathways, MDT use, and access to COCs. These findings support continued strengthening of screening participation, monitoring of care intervals, and quality assurance of MDT reporting and regional oncology care delivery.

## INTRODUCTION

Breast cancer is the most frequently diagnosed malignancy in women worldwide. In the Czech Republic, its burden has continued to rise: prevalence increased from approximately 1,300 cases per 100,000 women in 2012 to 1,800 per 100,000 in 2022, while incidence reached 7,564 newly diagnosed patients in 2022 [1].

The availability of modern targeted therapies, including immunotherapy, is very good in the Czech Republic and is covered by public health insurance. In the Czech Republic, oncology care is organized through a network of 16 Comprehensive Cancer Centres (COCs), 24 Regional Oncology Centres (ROCs), and Regional Oncology Groups across the country’s 14 regions [2]. COCs may delegate selected patients to ROCs, which are authorized to administer several centrally regulated systemic therapies used in breast cancer, including trastuzumab, bevacizumab, everolimus, and cyclin-dependent kinase inhibitors. Nevertheless, one region still lacks a COC, and three regions do not have an ROC. Only one centre (Masaryk Memorial Cancer Institute in Brno) currently meets the European standards for a comprehensive cancer centre according to the Organisation of European Cancer Institutes (OECI).

Mammography screening has transformed breast cancer into a disease that is both preventable through early intervention and detectable at an early stage [3-6]. In the Czech Republic, organized mammography screening has been in place since 2000 and is available to women aged 45 years and older, with no upper age limit. According to national data for 2024, 59.9 % of women aged 45–69 years underwent screening mammography within a two-year interval [7]. Screening is a key determinant of early-stage detection and favorable prognosis. In Czech women, 5-year relative survival decreases with advancing disease stage, from 100% in stage I patients to 93.6% and 73.9% in stage II and stage III patients, respectively [8].

Diagnostic mammography is a targeted breast imaging examination performed to evaluate a specific clinical problem, rather than as routine screening in women without symptoms. The mode of detection of breast cancer affects prognosis and may also shape the patient’s subsequent diagnostic and therapeutic pathway. Nationwide administrative and registry data make it possible to study such pathways at the population level. In several European countries, breast cancer patient pathways are an integral part of care organization and quality assurance, particularly within specialist breast centres [9], timed diagnostic pathways [10], and quality indicator-based monitoring systems [11-12].

The aim of this study was therefore to describe, both qualitatively and quantitatively, treatment pathways in women newly diagnosed with breast cancer in the Czech Republic, with a particular focus on comparing patients diagnosed through screening mammography (SCR) versus diagnostic mammography (DIG). The analyzed parameters included treatment centralization in COCs, first-line treatment (FLT), main treatment modality (MTM), time to treatment initiation, time to multidisciplinary team (MDT) discussion, and treatment outcomes according to disease stage and their temporal sequence. Analyses were performed for the whole country and further stratified by region and type of treatment center. Finally, quality indicators (QIs) were recommended to improve care for patients with breast cancer.

## METHODS

### Study design and data sources

This was a retrospective cohort study based on linked nationwide data from the Czech National Cancer Registry (NOR) and the National Registry of Reimbursed Health Services (NRHZS).

NOR contains information on oncological diseases, including date of diagnosis, provider of oncology care, and patient identifiers, and supports epidemiological analyses, health services research, and evaluation of preventive programs. NRHZS contains administrative data on reimbursed healthcare services, including the content and timing of services provided, healthcare provider, insured patient, and reimbursement information.

### Study population

Women were eligible if they had, in NOR, an invasive breast cancer (ICD-10 C50) as their first lifetime malignant neoplasm (excluding non-melanoma skin cancer, C44), diagnosed between 2017 and 2024, were aged ≥18 years on 1 January of the year of diagnosis, and female. Where a patient had more than one C50 tumor records, only the first primary C50 tumor was retained.

From this source population, a verified analytical cohort was defined by sequentially excluding patients with: a recorded death on or before the diagnosis date; missing administrative (NRHZS) records in the study window; internally inconsistent dates; no identifiable administrative index event (see below); treatment recorded before the index date; or an index date outside 2017–2024. The full attrition is shown in the cohort-selection diagram (Figure 1).

**Figure 1.**
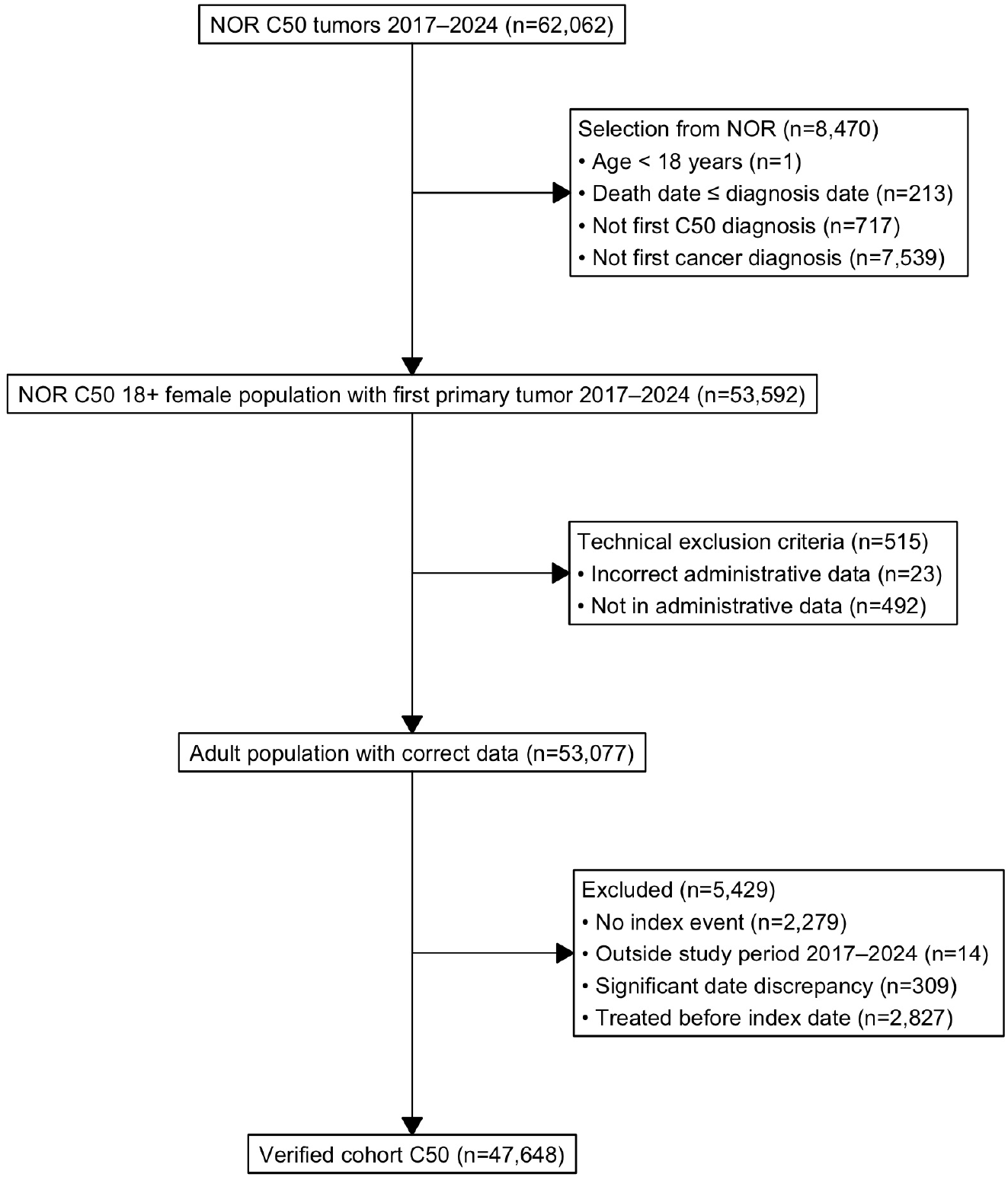
Overview of study design and patient cohort selection. *Abbreviations:* NOR, *National Cancer Registry;* C50, *Invasive breast cancer according the ICD-10 International Classification of Diseases, 10*^*th*^ *revision*.

### Definition of the administrative index event

The administrative index event was defined as the first mammography or breast ultrasound examination (screening or diagnostic) followed by histopathological examination (HP), subject to the following conditions: the interval between mammography or breast ultrasound and HP examination did not exceed 120 days; diagnosis code C50 was recorded within 150 days after mammography or breast ultrasound; diagnosis code C50 was recorded within 60 days after HP examination.

The date of mammography or breast ultrasound represented the index date. Patients without an identifiable imaging/HP combination were excluded from the verified analytical cohort. All time intervals reported here (time to treatment, time to MDT) are measured from this index date unless stated otherwise.

### Cohort and treatment-related entities definitions

Patients were classified as screen-detected (SCR) or diagnostically detected (DIG) according to the type (screening vs diagnostic) of the index mammography/ultrasound.

The analysis considered the following cohorts: verified cohort included patients verified by application of including and excluding criteria mentioned above; verified cohort detected by SCR mammography; verified cohort detected by DIG mammography; and verified treated cohort which included treated only patients.

First-line treatment (FLT) was defined as the first occurrence after the index date of surgery (SX), radiotherapy (RT) or systemic pharmacotherapy (PHT); patients with an FLT event were classified as treated and the remainder as untreated. The main treatment modality (MTM) was the principal modality delivered within one year after FLT, assigned by the priority SX > PHT > RT. Systemic therapy delivered within one year before surgery was considered neoadjuvant, and first-line treatment initially coded as systemic therapy in such patients was reclassified accordingly. Systemic-therapy subtypes (chemotherapy, endocrine therapy, targeted therapy and immunotherapy) were derived from ATC codes. Multidisciplinary team (MDT) discussion was ascertained from procedure codes recorded between the index date and FLT. Centralization of care was defined as delivery of FLT at a Comprehensive Cancer Centre (COC), identified from the national COC facility list, and was examined against the patient’s region of residence.

### Outcomes

The following outcomes were evaluated: age and year of diagnosis; stage distribution; proportion of untreated patients; FLT and MTM; time from index to treatment; time from index to MDT; centralization to COCs; and overall survival.

Overall survival (OS) was defined as all-cause survival from the index date to death from any cause, with patients alive censored at the administrative censoring date. The administrative censoring date was set to 31 December 2024, the last calendar period with complete mortality ascertainment in the linked data; person-time accruing afterwards was censored.

### Statistical analysis

All analyses were descriptive and exploratory, and no hypothesis testing or causal modelling was performed. Categorical variables are summarized as counts and percentages and continuous variables as means with standard deviations and medians with interquartile ranges. Survival was described using unadjusted Kaplan–Meier estimates by stage and detection mode, with numbers at risk. Because the comparisons between screen-detected and diagnostically detected patients are unadjusted, they should be read as associations rather than as effects of detection mode. Stage-stratified analyses were restricted to known stages (I–IV); cases with unknown/unclassifiable stage were excluded from those analyses and reported separately. Analyses were performed nationally and were further stratified by region and type of treatment centre. All analyses were conducted in R version 4.3.3.

## RESULTS

### Cohort characteristics

The verified cohort included 47,648 women with newly diagnosed breast cancer (Figure 1). Of these, 26,817 (56.3%) were classified as SCR and 20,831 (43.7%) as DIG. The median age was higher in SCR patients than in DIG patients (67 vs. 58 years).

### Stage at diagnosis

Stage distribution differed substantially between the two groups (Figure 2). In the SCR cohort, 17,498 (65.2%) of patients were diagnosed at stage I and 6,836 (25.5%) at stage II. In the DIG cohort, the corresponding proportions were 6,480 (31.1%) and 8,516 (40.9%), respectively. By contrast, advanced disease was more frequent among DIG patients: stage III accounted for 3,174 (15.2%) in DIG versus 1,219 (4.5%) in SCR, and stage IV for 1,891 (9.1%) in DIG versus 411 (1.5%) in SCR.

**Figure 2.**
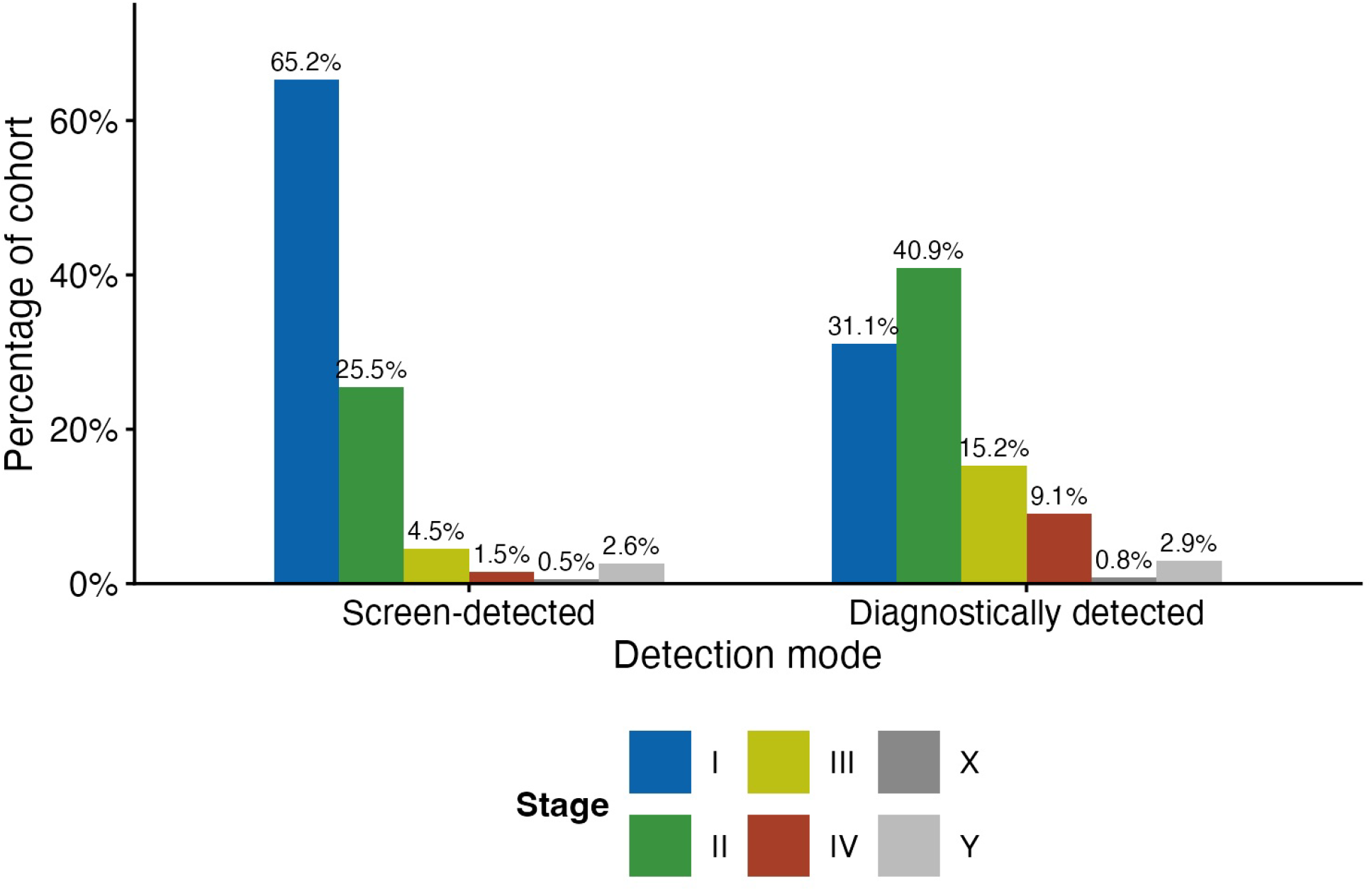
Stage distribution by detection mode in the verified cohort. *Abbreviations:* Stage X, *unknown due to objective limitations;* stage Y, *unknown due to missing data*.

### Untreated patients

Untreated patients constituted a very small subgroup: 483 of 47,648 women (1.0%). Of these, 388 belonged to the DIG group, meaning that 80.3% of untreated cases were diagnostically detected.

Median age among untreated DIG patients was 75 years, while untreated SCR patients had a median age of 72 years. In the untreated DIG group, 41.0% were diagnosed at stage IV and 48.2% died within 12 weeks after diagnosis, suggesting that non-treatment was concentrated among older, clinically fragile patients, particularly those with advanced disease (Supplementary Table S1).

### Treatment patterns

Surgery (SX) was the dominant MTM in stages I–III. However, the proportion of surgically treated patients decreased with advancing stage and was consistently lower in DIG than in SCR patients. By contrast, systemic treatment (PHT) became more frequent with increasing stage and was more common in the DIG cohort (Supplementary Figure S1).

In early breast cancer cases, FLT was mainly divided into SX and neoadjuvant therapy (Figure 3). In stage I, most women underwent SX as FLT, specifically 91.8% in SCR and 81.6% in DIG group. In stage II, the proportion of women receiving neoadjuvant therapy was nearly twice as high in DIG as in SCR patients (40.1% vs. 22.7%). In stage III, neoadjuvant therapy was administered to 46.6% of DIG patients and 40.9% of SCR patients.

**Figure 3.**
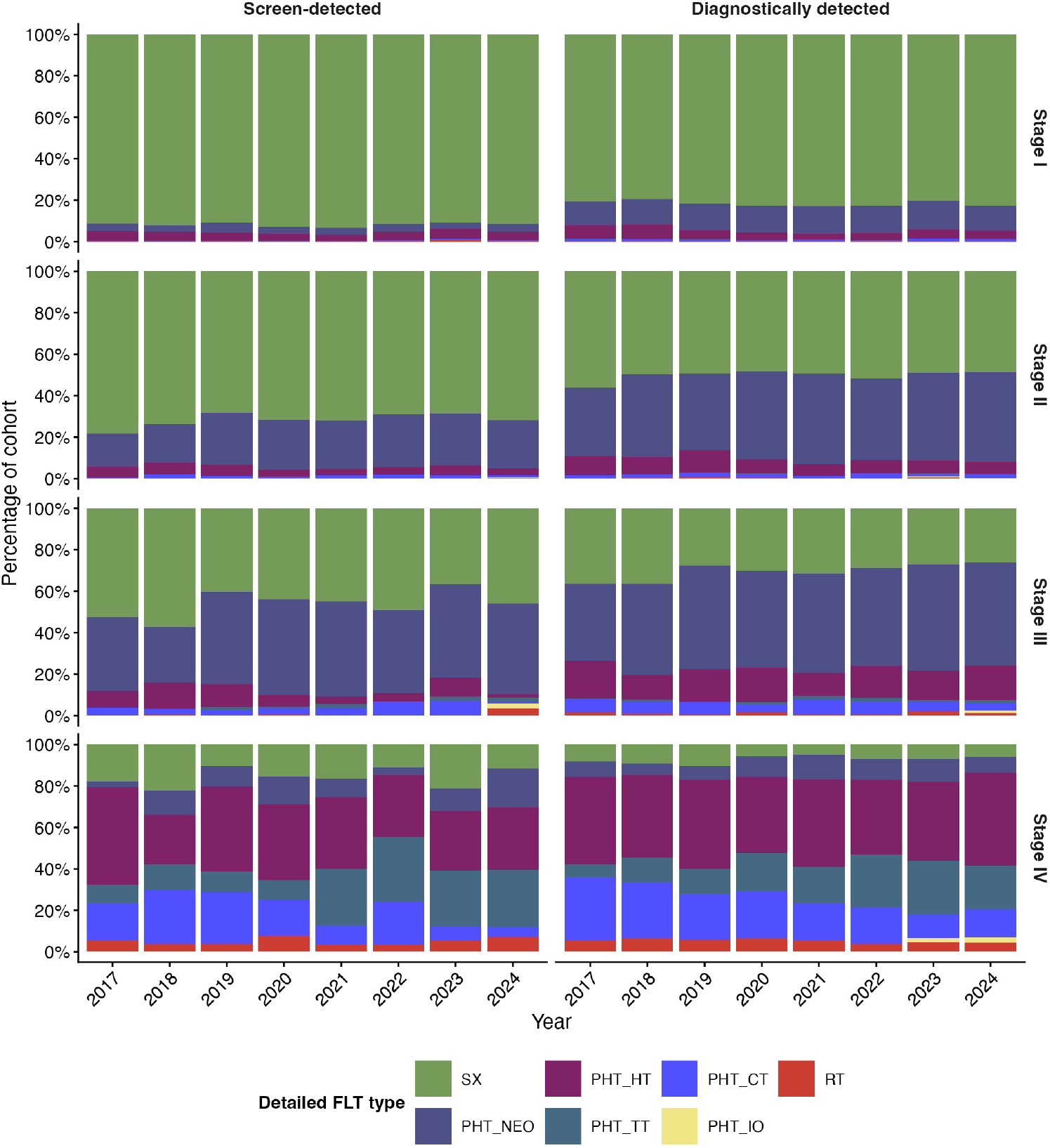
First treatment modalities by detection mode and disease stage across time. *Abbreviations:* FLT, *first-line treatment;* PHT, *pharmacotherapy;* HT, *hormonal therapy;* CT, *chemotherapy;* IO, *immunotherapy;* NEO, *neoadjuvant;* TT, *targeted therapy;* RT, *radiotherapy;* SX, *surgery*.

In stage IV, the use of chemotherapy as FLT decreased over time, whereas targeted therapy (PHT_TT) became more available after 2021.

### Time to treatment

Time to treatment was shorter among DIG patients than among SCR patients (Figure 4). In 2024, treatment within 4–6 weeks from mammography was achieved in 32.7% of SCR vs. 47.8% of DIG patients in stage I; 38.0% vs. 57.4% in stage II; 45.3% vs. 59.3% in stage III; and 44.3% vs. 63.3% in stage IV.

**Figure 4.**
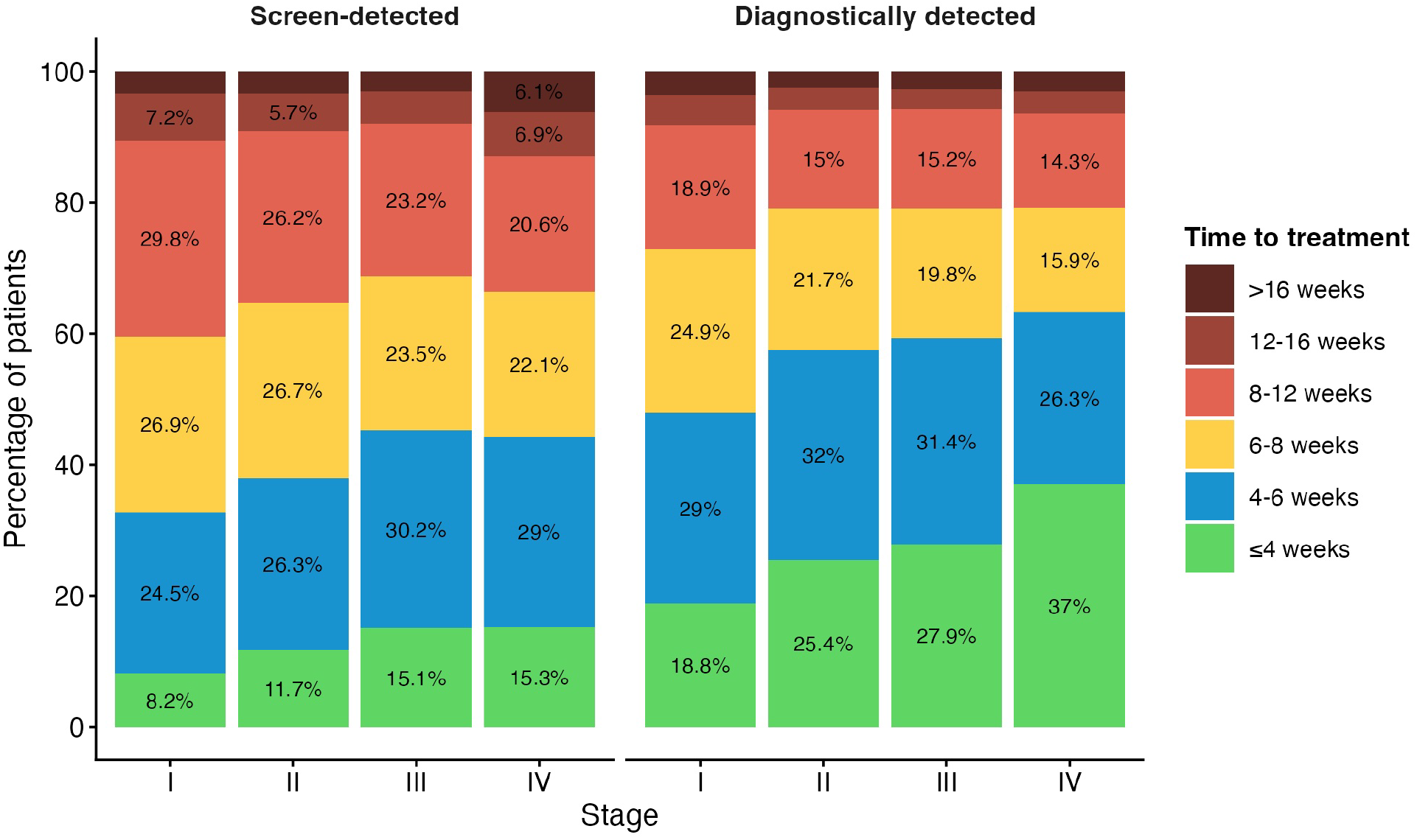
Time from index event to first-line treatment by detection mode and disease stage.

Conversely, treatment delays exceeding 8 weeks affected approximately one-third of SCR patients and one-quarter of DIG patients. Such delays were most frequent in stage I, where they occurred in 40.4% of SCR and 27.2% of DIG patients.

Regarding treatment modality, time to treatment was shorter for systemic PHT than for SX, likely reflecting the greater logistical complexity of operative planning. The median time to SX in stages I-III was 42-43 days in DIG patients, whereas in SCR patients it was consistently around 50 days. Median time to systemic therapy ranged from 33 to 35 days in DIG patients and from 40 to 46 days in SCR patients across stages I–IV. Detailed treatment intervals from the verified cohort are shown in Supplementary Table S2.

Marked regional differences (Figure 5) and differences between individual COCs (Supplementary Figure S2) were observed in time to treatment period.

**Figure 5.**
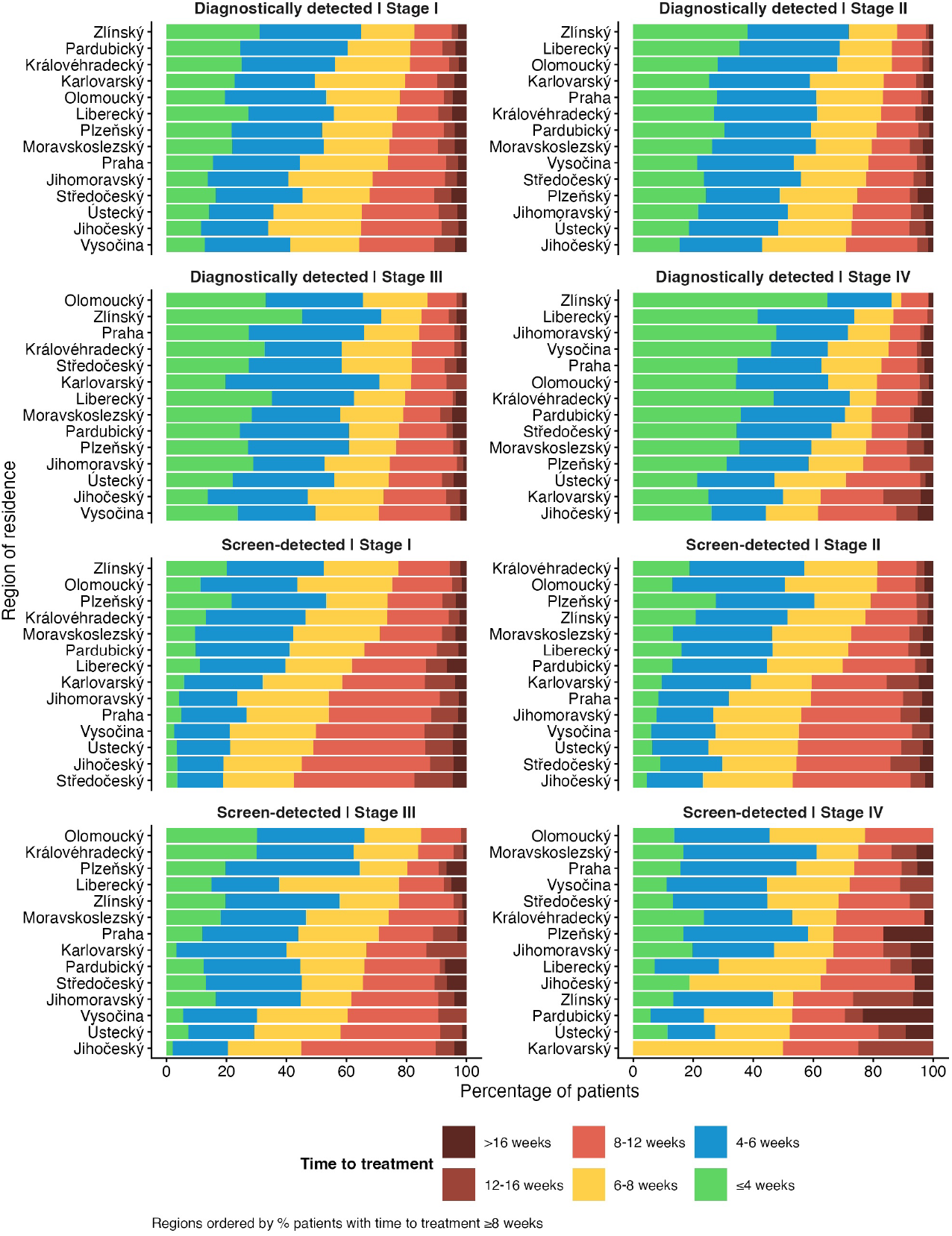
Time from index event to first-line treatment by detection mode and disease stage in different regions. Within each panel, regions are ordered by the percentage of patients with time to treatment ≥ 8 weeks.

### Multidisciplinary team involvement

The proportion of cases discussed by a MDT increased over time in both SCR and DIG groups. In 2024, MDT involvement reached approximately 82–86% in stages I–II and 71-82% in stages III-IV (Supplementary Figure S3).

Time from index date to MDT was longer in SCR than in DIG patients, particularly in stages II–III. For example, in 2024, MDT discussion within 4 weeks occurred in 69.1% of DIG patients versus 50.9% of SCR stage II patients and in 69.4% of DIG patients versus 55.1% of SCR stage III patients (Supplementary Table S3).

Substantial variability in MDT involvement and timing was observed across regions and different COCs.

### Centralization of care

The proportion of patients starting FLT in a COC increased over time, particularly in stages II–IV (Figure 6). In 2024, centralization reached 54.0% in stage I; 60.5% in stage II, 65.7% in stage III, and 74.1% in stage IV.

**Figure 6.**
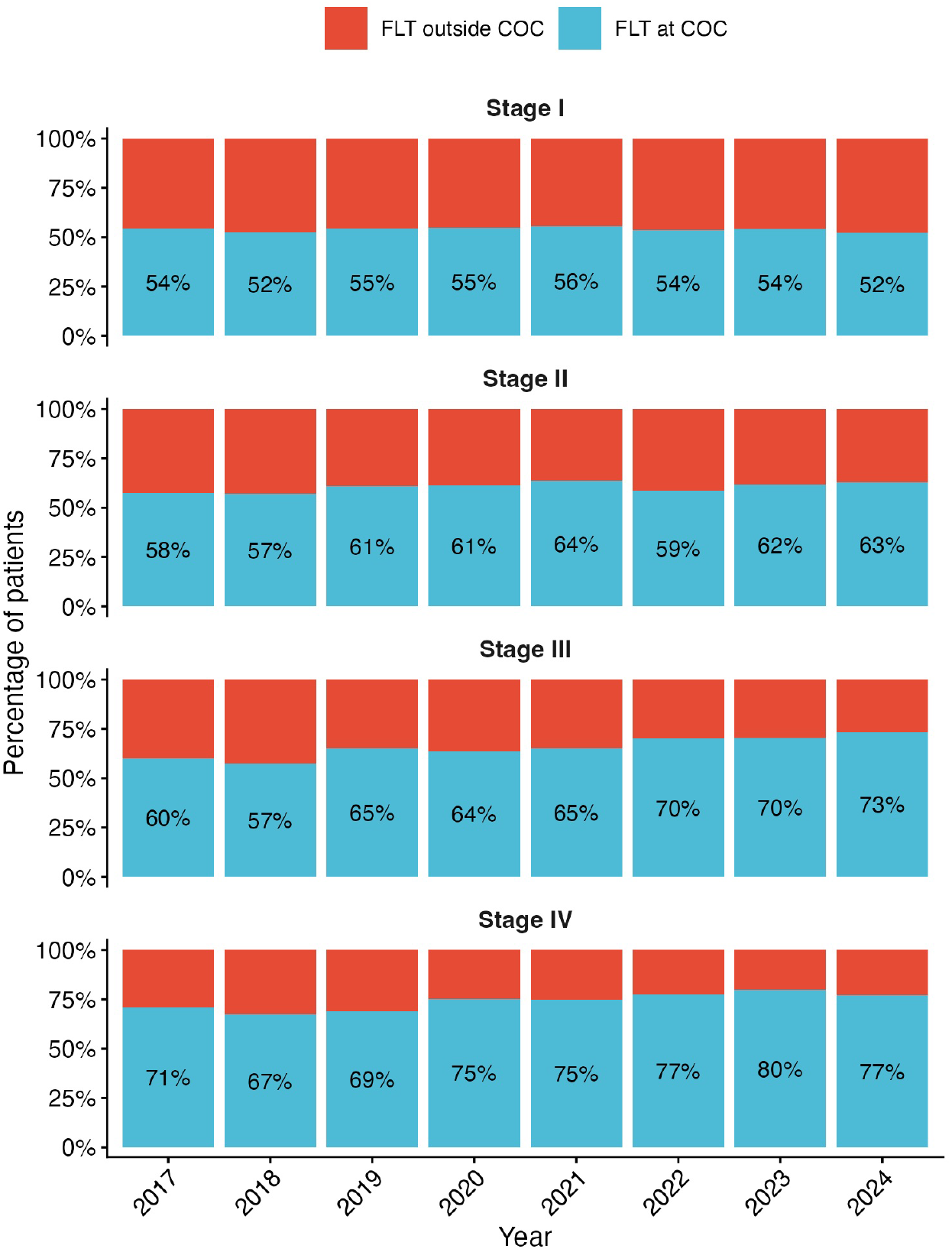
Centralization of first-line treatment to Comprehensive Cancer Centres by stage and year. *Abbreviations:* FLT, *first-line treatment;* COC, *Comprehensive Cancer Centre*.

Regional variation in care centralization was pronounced (Figure 7). Depending on the stage, the proportion of patients treated in COC ranged from 19.6% to 92.2% in stage I; 41.7% to 91.6% in stage II; 42.5% to 92.3% in stage III; and 50% to 97.8% in stage IV. These findings indicate substantial heterogeneity in access to specialized oncology care across the country.

**Figure 7.**
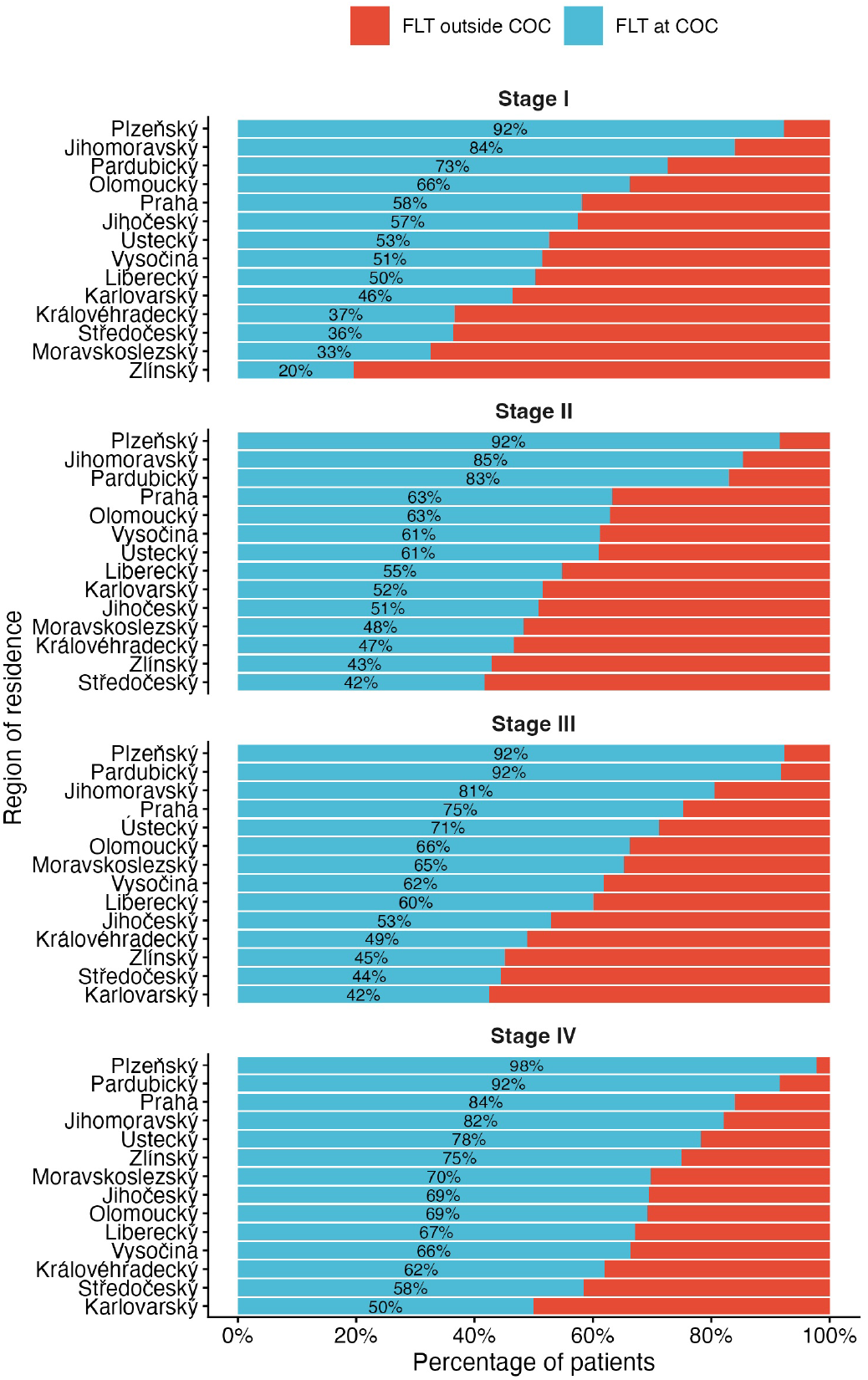
Centralization of first-line treatment to Comprehensive Cancer Centres by stage and year in different regions. *Abbreviations:* FLT, *first-line treatment;* COC, *Comprehensive Cancer Centre*.

### Survival

Survival patterns closely followed disease stage at diagnosis (Figure 8). Within the same stage, SCR patients showed more favorable survival curves than DIG patients. This difference likely reflects, at least in part, differences in case mix between the two groups, including age and biological aggressiveness not fully captured in the available data.

**Figure 8.**
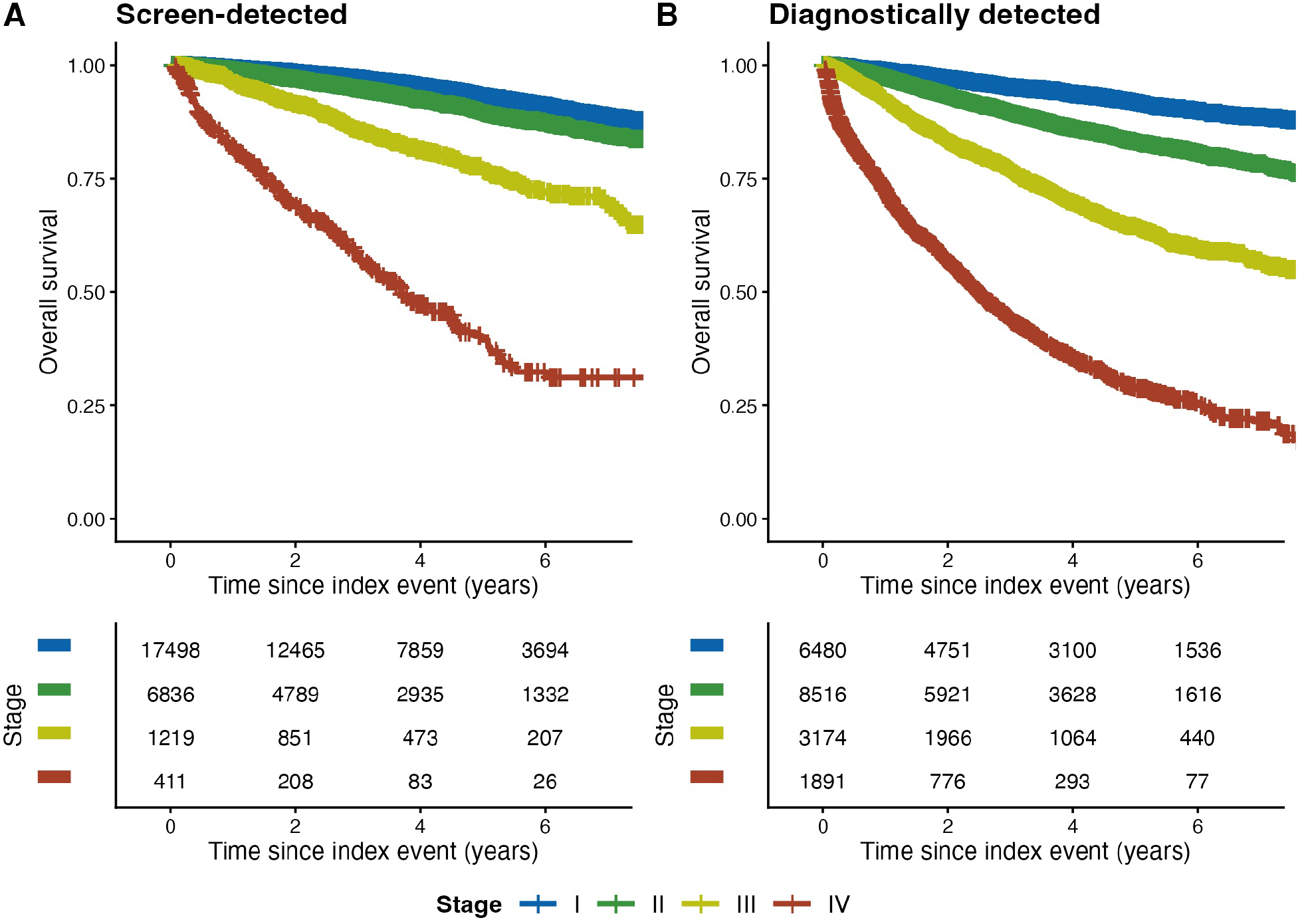
Survival of breast cancer patients according to disease stage and detection mode.

## DISCUSSION

Despite extensive evidence on the clinical benefits of mammographic screening, less is known about how the route of detection shapes subsequent diagnostic and treatment pathways at the population level. Our nationwide real-world study addresses this gap by providing a comprehensive description of breast cancer care pathways in the Czech Republic and demonstrating substantial differences between screening-detected and diagnostically detected cases.

The mode of detection was strongly associated with stage at diagnosis. Women diagnosed through SCR were much more likely to present with stage I disease, while DIG patients more frequently presented with advanced cancer. This pattern is consistent with the expected effect of organized mammography screening supporting its role in stage shift [13] and in about 20-25% reduction of cancer-specific mortality [4-6, 14]. Compared with other countries, breast cancer screening attendance in the Czech Republic (59.9% over the two-year period 2023-2024 [7]) is close to the EU average (∼57% in 2023) [15], but remains below the best-performing European programmes (∼80% in the Nordic countries) [16] and below reported screening uptake in the United States (∼80% in 2023) [17] and England (∼70% in 2023/2024) [18]. The present analysis further underscores the central role of screening in breast cancer care and supports the use of screening participation as a quality indicator at the population level.

In the Czech Republic, breast cancer screening starts relatively early, as the national program includes women from the age of 45 years [19]. This is earlier than the traditional approach used in many European countries, where organized screening has historically started at 50 years [20-21], but later than current U.S. recommendations, which advise biennial screening from 40 years [22]. Recent European Commission guidance also supports extending screening to women aged 45–74 years [23]. However, given the increasing incidence of breast cancer among young women in the Czech Republic — from 13 per 100,000 in 2000, to 19 per 100,000 in 2010, and 29 per 100,000 in 2020 — the question of lowering the starting age for screening below 45 years may deserve further consideration. Patients younger than 45 years represented 11% of the study cohort (n = 5,268 in total; mean 658 patients per year). This number is consistent with national data, according to which the absolute number of breast cancer cases among women younger than 45 years was approximately 400 in 2000, 570 in 2010, and 800 in 2020 [24].

Treatment rates of Czech breast cancer women were high; almost all women with stage I–III disease received active treatment. This contrasts with evidence from a large US cross-sectional study, which reported treatment refusal, with surgery being refused least often and chemotherapy most often [25]. However, racial and ethnic differences were described which are less common in the Czech Republic. In our cohort, untreated patients were predominantly older women with DIG-detected advanced disease and high short-term mortality, suggesting that non-treatment was concentrated among patients with poor clinical condition rather than reflecting limited access or low treatment willingness. Despite suboptimal participation in screening mammography, Czech women appear highly willing to undergo cancer treatment. This may be supported by broad treatment availability, good accessibility, and coverage by public health insurance. In older or more advanced patients, treatment type is particularly relevant, as many women with breast cancer remain eligible for well-tolerated endocrine or targeted therapy, thereby increasing the proportion of patients suitable for active treatment.

Treatment pathways differed systematically between SCR and DIG patients, indicating that these represent two groups of patients with different prognoses and different treatment requirements. DIG patients entered MDT discussion and initiated treatment faster than SCR patients. This difference was consistent across several stages and treatment modalities. It may reflect clinically appropriate prioritization of more advanced or symptomatic disease, which is in line with the relatively high accessibility of MDT discussion. A second explanation could be the concentration of patients in COCs; however, specialized care often only appear to shorten the diagnostic interval. The interval from diagnosis to treatment is not consistently shorter and may even be slightly longer in some population-based studies [26-27].

Our results also indicate that women diagnosed through screening may be at a disadvantage; treatment delays deserve attention, especially since screening is intended to improve outcomes through earlier diagnosis. The time to mammography was not measured directly; however, national data from mamo.cz indicate that the average waiting time for screening mammography is approximately 24 days, with a range of 13–70 days [28]. We observed substantial variability in time intervals among regions and individual COCs in the Czech Republic, which is likely consistent with the multifactorial causes of treatment delays. Published data also show heterogeneous results, with some studies reporting shorter [29], some similar [30], and others longer time to treatment in symptomatic patients compared with the screened population [31].

Although MDT coverage improved over time, it remained incomplete. Even in stages I–II, around one-quarter of patients were not captured as having undergone MDT discussion by 2022. Because the analysis relies on reimbursed procedure reporting, this may partly reflect variation in coding practices. Nevertheless, from a quality-of-care perspective, incomplete MDT reporting limits the ability to monitor adherence to recommended multidisciplinary management. Moreover, MDT discussion is associated with improved survival, and reduced recurrence, with observational studies suggesting approximately 10–20% better outcomes compared with non-MDT care [32-34].

Centralization of care in COCs increased over time, although substantial regional disparities remained. This was most apparent in patients with stage II–IV disease, many of whom require targeted therapies that are provided in COCs in the Czech Republic. In some regions, the proportion of patients treated in COCs remained below 50%; this may reflect differences in COC capacity, geographical access, availability of regional oncology services, referral organization, and patients’ willingness or ability to travel. In contrast, more than half of patients with stage I disease received care in COCs, although SX represents the standard primary treatment for cancers at this stage.

Centralization of surgical care is not finished yet in the Czech Republic; however a substantial number of breast cancer patients undergo SX in COCs. EUSOMA emphasized that breast cancer care should be centralized in specialist multidisciplinary breast centres and continuously monitored using standardized QIs to improve consistency, treatment quality, and patient outcomes [11]. Centralization of SX is associated with improved survival [35-36], higher adherence to guidelines [37-38], better surgical quality [39-40], broader access to multidisciplinary care [9, 11], and increased availability of breast-conserving and reconstructive procedures [41-42]. On the other hand, the centralization of oncology care faces capacity constraints that may affect the quality of cancer care, particularly time to treatment, which is influenced not only by waiting times for the first oncological consultation, but also by the limited capacity of imaging services.

We believe that high-quality care may be supported by a rational balance between centralization and decentralization. Decentralizing care for low-risk patients, selected high-risk patients during follow-up, frail patients requiring basic care, or patients unwilling to travel to specialist centres may release capacity in COCs. However, this process must be reversible, allowing patients to return to COCs when MDT discussion, specialized treatment, or advanced diagnostics are required. In the Czech Republic, patient transfer between COCs and regional hospitals is limited mainly by insufficient digitalization and patients’ reluctance to return to lower-level facilities. Care coordinators, whose position is now mandatory in all COCs, may help reduce unjustified variation between centres and maintain access to COC care for patients who need it most. Systematic measurement of care pathways and outcomes remains essential. Nationwide clinical auditing can improve breast cancer care through benchmarking, feedback, and monitoring of guideline adherence [12].

In conclusion, this nationwide retrospective cohort study showed that women with screen-detected breast cancer were diagnosed at earlier stages and had more favorable survival patterns than women diagnosed outside screening. At the same time, screen-detected patients experienced longer intervals to MDT discussion and treatment initiation. Treatment coverage in stages I–III was very high, MDT involvement increased over time, and centralization to COCs improved, particularly in more advanced stages. However, significant regional and center-level differences remained across the care pathway. These results support continued strengthening of mammography screening, monitoring of treatment timelines, improvement of MDT reporting and implementation, and targeted efforts to reduce regional disparities in access to specialized breast cancer care. To support these goals, several QIs were recommended for systematic measurement.

This study has several limitations. First, it is based on administrative and registry data and is therefore dependent on the completeness and validity of coding. In particular, MDT analyses were based on reported procedure codes and cannot distinguish true absence of MDT from underreporting. Second, the administrative definition of the index event, although operationally robust, may not capture all clinically relevant diagnostic trajectories. Cases without a detectable MMG-HP sequence were excluded from the verified cohort. Third, the study did not include detailed biological tumor characteristics, such as molecular subtype, HER2 status, hormone receptor status, or genomic risk stratification. Therefore, differences in treatment timing and modality may partly reflect unmeasured differences in tumor biology. Fourth, the analyses were exploratory and descriptive and were not designed to establish causal relationships. Finally, survival comparisons between SCR and DIG cohorts should be interpreted cautiously, as they may be influenced not only by stage distribution but also by lead-time bias, case-mix differences, and other unmeasured confounders. On the other hand, the analysis has several strengths, including the large number of patients analysed, good concordance with other national data, and the inclusion of variables — particularly time to treatment — that may help improve patient outcomes and reduce regional disparities in access to cancer care.

## Data Availability

Raw data can be legally required from the Institute of Health Information and STatistics of the Czech Republic

## DECLARATIONS

## Acknowledgments

The data were kindly provided by the Health Insurance Bureau in Prague, Czech Republic.

## Authors’ contributions

Conception/design: A.T., G.D., M.R. Investigation: G.D. Data analysis: A.T., G.D. Project administration: A.T. Data interpretation: A.T., G.D., Z.B., K.P., K.M., M.R., L.D., M.R. Manuscript writing: Z.B., G.D. Manuscript editing: all authors. Final approval of the manuscript: All authors.

## Ethics approval

This study was conducted using anonymized retrospective administrative claim data, and approval by an ethics committee was not required.

## Consent to Participate declaration

Not applicable.

## Data availability

The data remains property of the insurance companies and cannot be shared without their permission.

## Consent to Publish

All authors agree to the publication of this manuscript.

## SUPPLEMENTARY MATERIALS

**Figure S1**. Main treatment modality

**Figure S2**. Time to first-line treatment by facility

**Figure S3**. Multidisciplinary team assessment before treatment

**Table S1**. Baseline characteristics of untreated patients

**Table S2**. Time to first-line treatment

**Table S3**. Time to multidisciplinary team assessment

*Supplementary materials are provided as a separate file*.

## REFERENCES

1. Epidemiology of malignant breast neoplasm (C50) in women. Available at https://www.svod.cz/res/file/kartydiagnoz/c50.pdf

2. Official portal of the National Cancer Control Programme of the Czech Republic. Available at https://onconet.cz

3. Duffy, S. W., Tabár, L., Yen, A. M. F., Dean, P.B., Smith, R.A., et al. Mammography screening reduces rates of advanced and fatal breast cancers: results in 549,091 women. Cancer 126, 2971–2979 (2020). 10.1002/cncr.32859

4. Dibden, A., Offman, J., Duffy, S. W. & Gabe, R. Worldwide review and meta-analysis of cohort studies measuring the effect of mammography screening programmes on incidence-based breast cancer mortality. Cancers (Basel) 12, 976 (2020). 10.3390/cancers12040976

5. Nelson, H. D., Fu, R., Cantor, A., Pappas, M., Daeges, M., and Humphrey, L. Effectiveness of breast cancer screening: systematic review and meta-analysis to update the 2009 U.S. Preventive Services Task Force recommendation. Ann. Intern. Med. 164, 244–255 (2016). 10.7326/M15-0969

6. Lauby-Secretan, B., Scoccianti, C., Loomis, D., Benbrahim-Tallaa, L., Bouvard, V., Bianchini, F., and Straif, K. Breast-cancer screening — viewpoint of the IARC Working Group. N. Engl. J. Med. 372, 2353–2358 (2015). 10.1056/NEJMsr1504363

7. Chloupková, R., Ngo, O., Ambrožová, M., Hejcmanová, K., Dvořák, P., et al. Národní screeningové centrum: Datový portál screeningových programů. Ministerstvo zdravotnictví ČR, Praha (2022). Available at: https://nscdata.uzis.cz/cs/data/screening-karcinom-prsu/interaktivni-vizualizace/mamo-pokryti/ Accessed 21 June 2026.

8. Krejčí, D., Mužík, J., Šnábl, I., Gregor, J., Komenda, M., Dušek, L. Portal of Cancer Epidemiology in the Czech Republic. Available at: https://www.svod.cz/preziti Accessed 21 June 2026.

9. Wilson, A. R. M., Marotti, L., Bianchi, S., Biganzoli, L., Claassen, S., Decker, T. et al. The requirements of a specialist Breast Centre. Eur. J. Cancer 49, 3579–3587 (2013). 10.1016/j.ejca.2013.07.017

10. Héquet, D., Huchon, C., Baffert, S., Alran, S., Reyal, F., Nguyen, T. et al. Preoperative clinical pathway of breast cancer patients: determinants of compliance with EUSOMA quality indicators. Br. J. Cancer 116, 1394–1401 (2017). 10.1038/bjc.2017.114

11. Biganzoli, L., Marotti, L., Hart, C. D., Cataliotti, L., Cutuli, B., Kühn, T. et al. Quality indicators in breast cancer care: an update from the EUSOMA working group. Eur. J. Cancer 86, 59–81 (2017). 10.1016/j.ejca.2017.08.017

12. van Bommel, A. C. M., Spronk, P. E. R., Vrancken Peeters, M. T. F. D., Jager, A., Lobbes, M., Maduro, J.H., et al. Clinical auditing as an instrument for quality improvement in breast cancer care in the Netherlands: the national NABON Breast Cancer Audit. J. Surg. Oncol. 115, 243–249 (2017). 10.1002/jso.24516

13. de Munck, L., Siesling, S., Fracheboud, J., Heeten, G.J., Broeders, M.J., de Bock, H.J. Impact of mammographic screening and advanced cancer definition on the percentage of advanced-stage cancers in a steady-state breast screening programme in the Netherlands. Br. J. Cancer 123, 1191–1197 (2020). 10.1038/s41416-020-0968-6

14. Zielonke, N., Gini, A., Jansen, E. E. L., Antilla, A., Segnan, N., Ponti, A., et al. Evidence for reducing cancer-specific mortality due to screening for breast cancer in Europe: a systematic review. Eur. J. Cancer 127, 191–206 (2020). 10.1016/j.ejca.2019.12.010

15. European Commission. European Cancer Information System (ECIS): Breast cancer screening coverage in EU countries. European Commission, Brussels (2023). Available at: https://ecis.jrc.ec.europa.eu

16. OECD/European Union. Health at a Glance: Europe 2024. State of Health in the EU Cycle. OECD Publishing, Paris (2024). Available at: https://www.oecd.org/health/health-at-a-glance-europe/

17. Centers for Disease Control and Prevention. Mammography use among women aged 50–74 years — United States, 2023. Prev. Chronic Dis. 22, 250139 (2025). Available at: https://www.cdc.gov/pcd/issues/2025/25_0139.htm

18. NHS England. Breast screening programme, England: statistics for 2023 to 2024. NHS England, London (2024). Available at: https://digital.nhs.uk/data-and-information/publications/statistical/breast-screening-programme

19. Czech Breast Cancer Screening Programme. About the programme. Available at: https://www.mamo.cz/en/about/

20. OECD. Health at a Glance 2025: Cancer screening. Organisation for Economic Co-operation and Development, Paris (2025). Available at: https://www.oecd.org/en/publications/health-at-a-glance-2025_8f9e3f98-en/full-report/cancer-screening_a3f047dd.html

21. NHS England. NHS Breast Screening Programme: Helping you decide. Available at: https://www.gov.uk/government/publications/breast-screening-helping-women-decide/nhs-breast-screening-helping-you-decide

22. US Preventive Services Task Force. Screening for breast cancer: US Preventive Services Task Force recommendation statement. JAMA 331, 1918–1930 (2024). 10.1001/jama.2024.5534

23. European Commission. Proposal for a Council Recommendation on strengthening prevention through early detection: a new EU approach on cancer screening replacing Council Recommendation 2003/878/EC. European Commission, Brussels (2022). Available at: https://health.ec.europa.eu/system/files/2022-09/com_2022-474_act_en.pdf

24. Krejčí, D., Mužík, J., Šnábl, I., Gregor, J., Komenda, M., Dušek, L. Portal of Cancer Epidemiology in the Czech Republic. Available at: https://www.svod.cz/incidence (Accessed 21 June 2026).

25. Freeman, J.Q., Li, J.L., Fisher, S.G., Yao, K.A., David, S.P., Huo, D. Declination of Treatment, Racial and Ethnic Disparity, and Overall Survival in US Patients With Breast Cancer. JAMA Netw. Open 2024 May 1;7(5):e249449. 10.1001/jamanetworkopen.2024.9449

26. Webber, C., Jiang, L., Grunfeld, E. & Groome, P. A. Breast cancer diagnosis and treatment wait times in specialized diagnostic units compared with usual care: a population-based study. Curr. Oncol. 27, e546–e553 (2020). 10.3747/co.27.6105

27. Chiarelli, A. M., Blackmore, K. M., Muradali, D., Smith, C.R., Mirea, L., Majpruy, V., et al. Evaluating wait times from screening to breast cancer diagnosis among women undergoing organised assessment vs usual care. Br. J. Cancer 116, 1254–1263 (2017). 10.1038/bjc.2017.87

28. Plotogea, A. D., Chiarelli, A. M., Mirea, L., Prummel, M. V., Chong, N. & Shumak, R. S. et al. Clinical and prognostic factors associated with diagnostic wait times by breast cancer detection method. SpringerPlus 3, 125 (2014). 10.1186/2193-1801-3-125

29. Yuan, Y., Li, M., Yang, J., Elliot, T., Dabbs, K., Dickinson, J. A. et al. The diagnostic interval for breast cancer in Northern Alberta: do screen-detected and symptom-detected cancers differ? BMC Health Serv. Res. 16, 684 (2016). 10.1186/s12913-016-1303-z

30. Jiang, L., Gilbert, J., Langley, H., Moineddin, R. & Groome, P. A. Regional variation in diagnostic intervals for breast cancer in Ontario and the effect of detection method and assessment units. Health Promot. Chronic Dis. Prev. Can. 38, 367–376 (2018). 10.24095/hpcdp.38.10.02

31. Kesson, E. M., Allardice, G. M., George, W. D., Burns, H. J. G. & Morrison, D. S. Effects of multidisciplinary team working on breast cancer survival: retrospective, comparative, interventional cohort study of 13,722 women. BMJ 344, e2718 (2012). 10.1136/bmj.e2718

32. Tsai, C. H., Hsieh, H. F., Lai, T. W., Kung, P. T., Kuo, W. Y. & Tsai, W. C. Effect of multidisciplinary team care on the risk of recurrence in breast cancer patients: a national matched cohort study. Breast 53, 68–76 (2020). 10.1016/j.breast.2020.07.008

33. Coory, M., Gkolia, P., Yang, I. A., Bowman, R. V. & Fong, K. M. Systematic review of multidisciplinary teams in the management of lung cancer. Lung Cancer 60, 14–21 (2008). 10.1016/j.lungcan.2007.09.004

34. Gooiker, G. A., van Gijn, W., Wouters, M. W. J. M., Post, P. N., van de Velde, C. J. H. & Tollenaar, R. A. E. M. A systematic review and meta-analysis of the volume-outcome relationship in the surgical treatment of breast cancer. Eur. J. Surg. Oncol. 36 Suppl. 1, S27–S35 (2010). 10.1016/j.ejso.2010.06.024

35. Greenup, R. A., Obeng-Gyasi, S., Thomas, S., Houck, K., Lane, W. O. & Blitzblau, R. C. et al. The effect of hospital volume on breast cancer mortality. Ann. Surg. 267, 375–381 (2018). 10.1097/SLA.0000000000002095

36. Kreienberg, R., Wöckel, A. & Wischnewsky, M. Highly significant improvement in guideline adherence, relapse-free and overall survival in breast cancer patients when treated at certified breast cancer centres: an evaluation of 8323 patients. Breast 40, 54–59 (2018). 10.1016/j.breast.2018.04.002

37. Ricci-Cabello, I., Saletti-Cuesta, L., Slight, S. P. & Valderas, J. M. Adherence to breast cancer guidelines is associated with better survival outcomes: a systematic review and meta-analysis of observational studies in EU countries. BMC Health Serv. Res. 20, 920 (2020). 10.1186/s12913-020-05749-x

38. Jeevan, R., Cromwell, D. A., Trivella, M., Lawrence, G., Kearins, O. & Pereira, J. et al. Reoperation rates after breast conserving surgery for breast cancer among women in England: retrospective study of hospital episode statistics. BMJ 345, e4505 (2012). 10.1136/bmj.e4505

39. Tamburelli, F., Maggiorotto, F., Marchiò, C., Balmativola, D., Magistris, A. & Kubatzki, F. et al. Reoperation rate after breast conserving surgery as quality indicator in breast cancer treatment. Breast Cancer 27, 829–836 (2020). 10.1007/s12282-020-01083-8

40. de Boniface, J., Szulkin, R. & Johansson, A. L. V. Survival after breast conservation vs mastectomy adjusted for comorbidity and socioeconomic status: a Swedish national 6-year follow-up of 48,986 women. JAMA Surg. 156, 628–637 (2021). 10.1001/jamasurg.2021.1438

41. Nicholson, K., Kuchta, K., Bleicher, R., Stevens, R., Dietz, J. & Wilke, L. et al. Impact of the National Accreditation Program for Breast Centers reconstruction standard on reconstruction rates at Commission on Cancer centers with breast center accreditation. Ann. Breast Surg. 8, 14 (2024). 10.21037/abs-23-3

