## Supplementary for "Screen-Detected and Diagnostic Breast Cancers Show Distinct Treatment Pathways and Quality Indicator Performance"

### Supplementary Material

#### Contents

Figure S1. Main treatment modality
Figure S2. Time to first-line treatment by facility
Figure S3. Multidisciplinary team assessment before treatment
Table S1. Baseline characteristics of untreated patients
Table S2. Time to first-line treatment
Table S3. Time to multidisciplinary team assessment

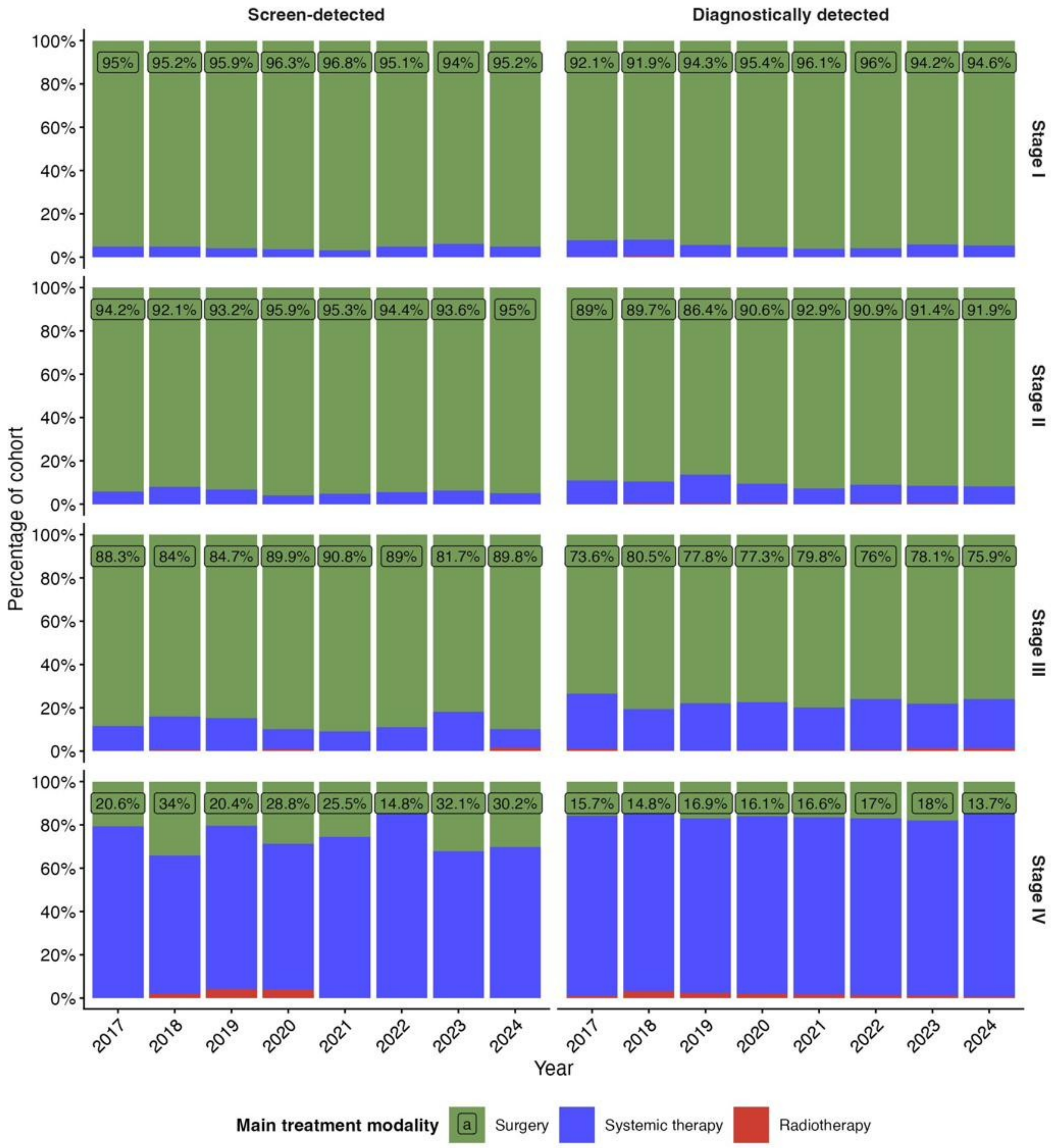


**Figure S1.** Main treatment modality in the verified treated cohort, by disease stage, detection mode and year of diagnosis. The value labeled on each bar is the proportion of patients treated with surgery.

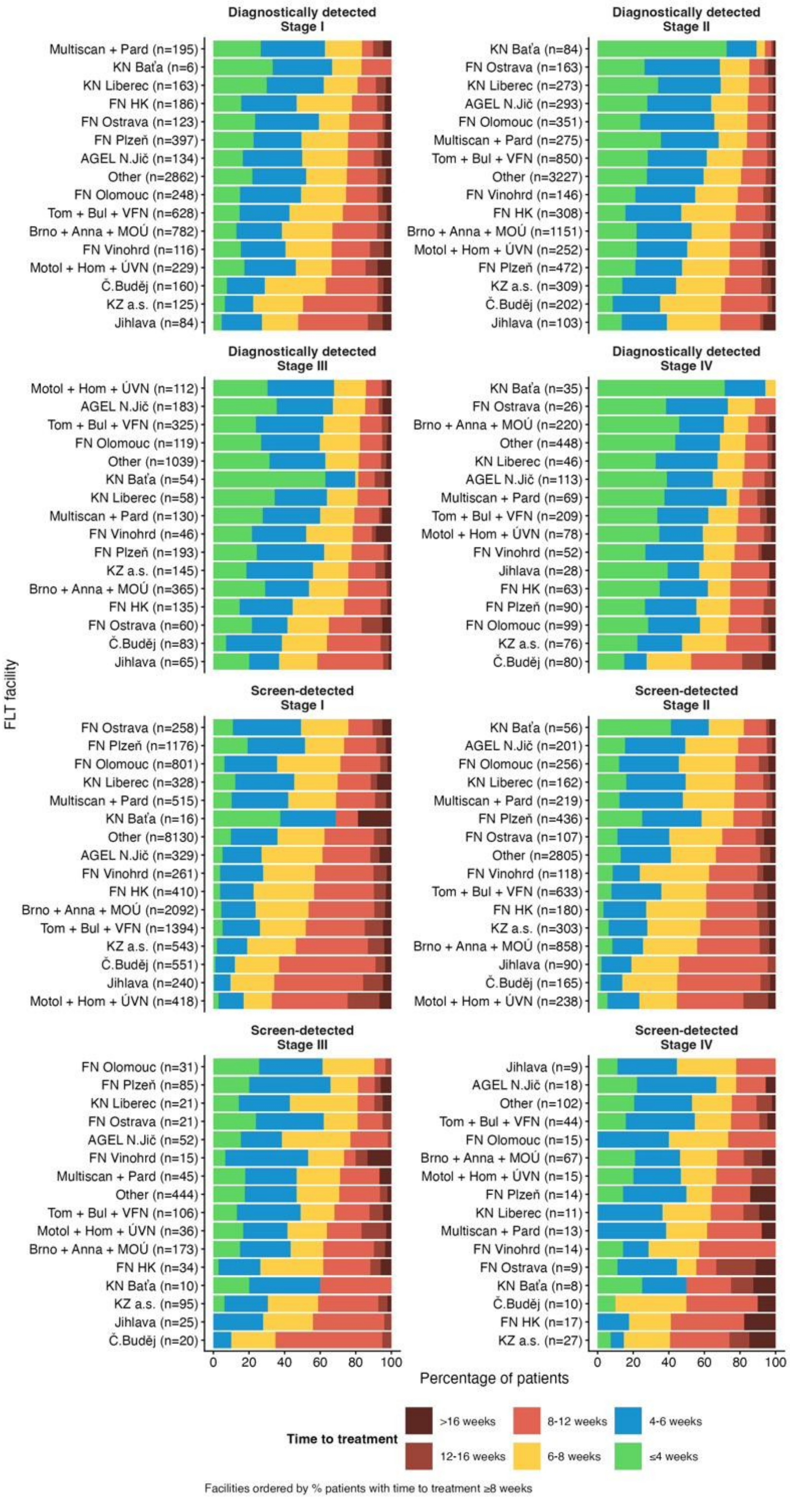


**Figure S2.** Time from index event to first-line treatment, by disease stage, detection mode and treating facility (n shown per facility). Within each panel, facilities are ordered by the percentage of patients with time to treatment ≥ 8 weeks.

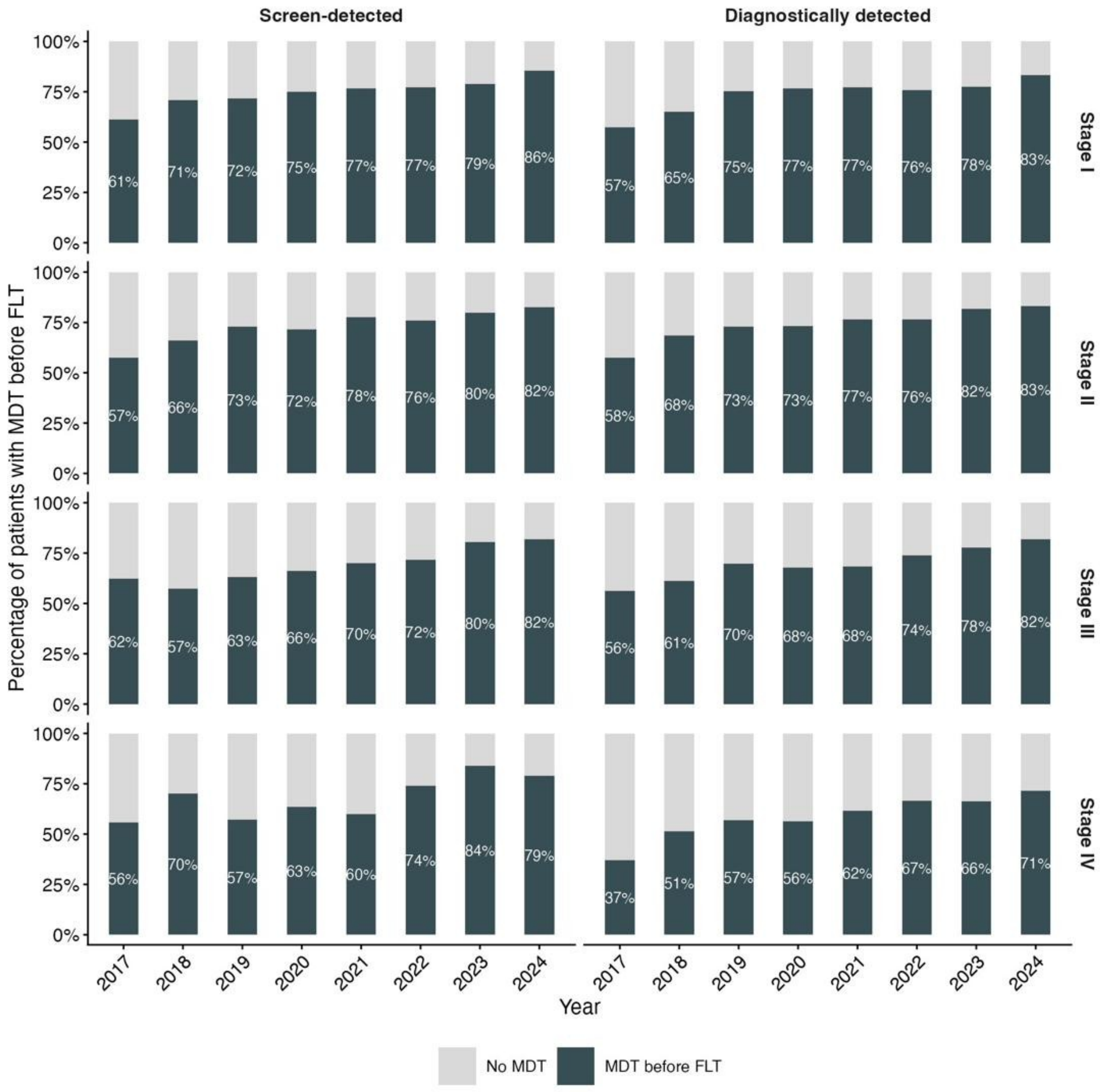


**Figure S3.** Proportion of patients with a multidisciplinary team (MDT) assessment before first-line treatment, by disease stage, detection mode and year of diagnosis.

**Table S1.** Baseline characteristics of untreated patients in the verified cohort, by detection mode and disease stage. Data are mean (SD), median (Q1, Q3) or n (%).

| Characteristic | Screen-detected (N = 69) | | | | | Diagnostically detected (N = 320) | | | | |
|---|---|---|---|---|---|---|---|---|---|---|
| | **Total** n=69 | **I** n=36 | **II** n=9 | **III** n=6 | **IV** n=18 | **Total** n=320 | **I** n=42 | **II** n=57 | **III** n=62 | **IV** n=159 |
| **Age (years)** | | | | | | | | | | |
| Mean (SD) | 69 (13) | 67 (13) | 68 (17) | 76 (13) | 74 (11) | 71 (15) | 71 (19) | 75 (16) | 76 (14) | 68 (13) |
| Median (Q1, Q3) | 71 (59, 81) | 69 (55, 77) | 70 (53, 83) | 78 (63, 81) | 78 (64, 81) | 75 (60, 83) | 77 (55, 86) | 79 (67, 89) | 82 (67, 86) | 71 (59, 78) |
| **Year of diagnosis** | | | | | | | | | | |
| 2017 | 11 (16%) | 6 (17%) | 2 (22%) | 1 (17%) | 2 (11%) | 41 (13%) | 2 (4.8%) | 7 (12%) | 6 (9.7%) | 26 (16%) |
| 2018 | 7 (10%) | 3 (8.3%) | 1 (11%) | 2 (33%) | 1 (5.6%) | 41 (13%) | 9 (21%) | 5 (8.8%) | 7 (11%) | 20 (13%) |
| 2019 | 12 (17%) | 7 (19%) | 1 (11%) | 2 (33%) | 2 (11%) | 45 (14%) | 8 (19%) | 12 (21%) | 8 (13%) | 17 (11%) |
| 2020 | 9 (13%) | 5 (14%) | 0 (0%) | 0 (0%) | 4 (22%) | 34 (11%) | 2 (4.8%) | 9 (16%) | 6 (9.7%) | 17 (11%) |
| 2021 | 4 (5.8%) | 3 (8.3%) | 0 (0%) | 1 (17%) | 0 (0%) | 27 (8.4%) | 3 (7.1%) | 4 (7.0%) | 5 (8.1%) | 15 (9.4%) |
| 2022 | 7 (10%) | 3 (8.3%) | 1 (11%) | 0 (0%) | 3 (17%) | 47 (15%) | 3 (7.1%) | 7 (12%) | 8 (13%) | 29 (18%) |
| 2023 | 13 (19%) | 5 (14%) | 4 (44%) | 0 (0%) | 4 (22%) | 40 (13%) | 8 (19%) | 6 (11%) | 8 (13%) | 18 (11%) |
| 2024 | 6 (8.7%) | 4 (11%) | 0 (0%) | 0 (0%) | 2 (11%) | 45 (14%) | 7 (17%) | 7 (12%) | 14 (23%) | 17 (11%) |
| **Time to death** | | | | | | | | | | |
| ≤4 weeks | 3 (4.3%) | 0 (0%) | 1 (11%) | 0 (0%) | 2 (11%) | 63 (20%) | 4 (9.5%) | 5 (8.8%) | 5 (8.1%) | 49 (31%) |
| 5–8 weeks | 6 (8.7%) | 0 (0%) | 1 (11%) | 1 (17%) | 4 (22%) | 71 (22%) | 4 (9.5%) | 11 (19%) | 9 (15%) | 47 (30%) |
| 9–12 weeks | 3 (4.3%) | 2 (5.6%) | 0 (0%) | 0 (0%) | 1 (5.6%) | 22 (6.9%) | 1 (2.4%) | 1 (1.8%) | 3 (4.8%) | 17 (11%) |
| >12 weeks | 27 (39%) | 10 (28%) | 2 (22%) | 5 (83%) | 10 (56%) | 112 (35%) | 16 (38%) | 25 (44%) | 33 (53%) | 38 (24%) |
| Censored (alive) | 30 (43%) | 24 (67%) | 5 (56%) | 0 (0%) | 1 (5.6%) | 52 (16%) | 17 (40%) | 15 (26%) | 12 (19%) | 8 (5.0%) |

**Table S2.** Time from index event to first-line treatment (days) in the verified cohort, by detection mode, disease stage and treatment type.

| Stage / treatment | Screen-detected | | | | Diagnostically detected | | | |
|---|---|---|---|---|---|---|---|---|
| | N | Mean | 80th pct | Median | N | Mean | 80th pct | Median |
| **Stage I** | | | | | | | | |
| Surgery | 16,025 | 54 | 69 | 50 | 5,252 | 49 | 62 | 43 |
| Systemic therapy | 1,384 | 57 | 82 | 46 | 1,166 | 46 | 56 | 35 |
| Radiotherapy | 53 | 134 | 152 | 105 | 20 | 169 | 139 | 118 |
| **Stage II** | | | | | | | | |
| Surgery | 4,882 | 54 | 70 | 50 | 4,253 | 47 | 62 | 43 |
| Systemic therapy | 1,934 | 47 | 59 | 40 | 4,173 | 40 | 49 | 33 |
| Radiotherapy | 11 | 144 | 120 | 91 | 33 | 107 | 115 | 92 |
| **Stage III** | | | | | | | | |
| Surgery | 563 | 52 | 69 | 49 | 950 | 48 | 63 | 44 |
| Systemic therapy | 643 | 51 | 59 | 40 | 2,126 | 44 | 51 | 34 |
| Radiotherapy | 7 | 101 | 129 | 83 | 36 | 63 | 78 | 56 |
| **Stage IV** | | | | | | | | |
| Surgery | 62 | 55 | 69 | 49 | 124 | 45 | 56 | 40 |
| Systemic therapy | 311 | 66 | 69 | 43 | 1,515 | 47 | 56 | 34 |
| Radiotherapy | 20 | 51 | 82 | 42 | 93 | 47 | 51 | 29 |

**Table S3.** Time from index event to multidisciplinary team (MDT) assessment in the verified treated cohort, by detection mode and disease stage. Values are the percentage of patients within each interval.

| Stage | <2 weeks | 2–4 weeks | 4–6 weeks | >6 weeks |
|---|---|---|---|---|
| **Screen-detected** | | | | |
| I | 10.0 | 39.9 | 27.8 | 22.3 |
| II | 10.8 | 40.1 | 27.6 | 21.5 |
| III | 11.1 | 44.0 | 26.1 | 18.7 |
| IV | 17.5 | 39.0 | 25.7 | 17.8 |
| **Diagnostically detected** | | | | |
| I | 20.2 | 44.1 | 21.0 | 14.8 |
| II | 22.4 | 46.7 | 19.6 | 11.4 |
| III | 25.6 | 43.9 | 19.1 | 11.5 |
| IV | 28.9 | 45.1 | 15.9 | 10.2 |
